# Characteristic resting state facial expressions in older adults with mild cognitive impairment level

**DOI:** 10.64898/2026.04.10.26350581

**Authors:** Mirano Miyayama, Takuya Sekiguchi, Hikaru Sugimoto, Toshikazu Kawagoe, Kornkanok Tripanpitak, Alexandra Wolf, Kazumi Kumagai, Kosuke Fukumori, Kumi Watanabe Miura, Shogo Okada, Keiichiro Ishimaru, Mihoko Otake-Matsuura

## Abstract

**Background:** For early detection of Alzheimer’s disease, it is essential to identify individuals showing cognitive performance consistent with the mild cognitive impairment (MCI) range during preliminary screening, ideally using methods that extend beyond conventional cognitive assessments. Non-invasive, easily accessible screening tools applicable in daily life are increasingly needed. Facial expressions, particularly during rest, may offer promising biomarkers for MCI level detection. This study aimed to identify specific facial features associated with MCI level during rest to inform development of facial expression-based screening tools.

**Methods:** Participants were classified into an MCI level group and a healthy control (HC) group based on the Montreal Cognitive Assessment (MoCA) scores. Facial Action Units (AUs) were extracted from video recordings of resting-state facial expressions in 31 individuals with MCI level and 14 HC. Two statistical models were employed: a multilevel zero-inflated beta regression model for intensity of 17 AUs and a multilevel logistic regression model for presence or absence of 18 AUs.

**Results:** In the zero-inflated beta regression, the AU relates to upper lip raiser showed a significant group effect (MCI level vs. HC; *p* <0.001), remaining significant after multiple comparison correction. The logistic regression revealed significant group differences for the AUs related to lip tightener (*p* <0.001) and lip suck (*p* <0.001), both remained significant after multiple comparison correction.

**Conclusions:** Distinctive facial action patterns during rest were observed in individuals with MCI level. These findings highlight the potential of resting-state facial expressions as a basis for novel, unobtrusive screening tools for early MCI level detection.

## Background

Mild cognitive impairment (MCI) is a prodromal stage of dementia characterized by cognitive decline that cannot be attributed to normal aging. However, it does not meet the diagnostic criteria for dementia, and individuals with MCI typically maintain their activities of daily living ^1^. Despite preserved daily functioning, the extent of brain lesions in this population is intermediate between that of healthy individuals and dementia patients ^2^.

Some people with MCI convert to dementia, with annual rates of around 10% ^3^. As populations around the world continue to age, the prevalence of MCI is becoming an increasingly critical public health issue. In Japan alone, the number of individuals with MCI is estimated to be around 5.58 million^4^, underscoring the urgency of addressing this challenge.

Despite these observations, a proportion of individuals with MCI may revert to normal cognitive function ^5^. For instance, Chung et al. ^6^ reported that 39% of individuals with MCI returned to a cognitively healthy state. In the absence of curative treatments for dementia, early detection and timely intervention at the MCI stage remain essential ^7,8^.

The early detection of MCI remains challenging ^9^. One contributing factor is that current methods for detecting MCI are not easily accessible, as they often involve procedures such as neuropsychological assessments and neuroimaging conducted in clinical settings ^10^, which may impose a burden on patients. In addition, because MCI typically does not interfere with activities of daily living, individuals may lack awareness of their condition ^9^. These limitations highlight the need for more accessible and less stressful MCI screening tools that can be implemented in everyday settings.

One possible new screening method for MCI is the use of face data. Facial expressions are considered important information in clinical psychiatry as an adjunct to diagnosis ^11^. Objectively quantifying facial features used to be difficult, but recent technological advancements have made this more possible, and characteristic facial expressions specific to mental disorders have been the focus of several recent research studies ^12,13^. For example, Jung et al. ^14^ found that there are characteristic facial expressions, such as lip press and stretch were attenuated in depressive patients compared to those in healthy controls (HC).

Characteristic facial expressions have been reported not only in mental disorders, but also in dementia. For example, in Alzheimer’s Disease (AD) patients, when they view a sequence of emotion-eliciting and neutral images, it has been noted that their degree of cognitive decline corresponds to increased facial expressiveness ^15^. Additionally, Burton and Kaszniak ^16^ reported that, while viewing emotion-eliciting images, zygomatic activity was different between AD and control groups. It has also been reported that artificial intelligence (AI), deep learning, or machine learning approach can distinguish between dementia and healthy groups based on video-recordings of facial expressions during conversations and when participants are passively viewing images ^17–20^. Particularly when examining facial images with neutral expressions, deep learning model classified dementia and HC with a high degree of accuracy (area under the curve: 0.925) ^21^. Based on these findings, it is suggested that patients with dementia exhibit characteristic facial expressions, particularly in neutral facial expressions.

However, studies using faces with neutral expressions have limitations, due to the complexity and black-box nature of AI systems, which make it unclear specifically which facial features are used for classification ^18,21^. Considering that certain facial muscles are active during the resting state ^22^, AI systems may make their classifications based on a specific way of holding the face—that is, the way the facial muscles are used, as reflected in the appearance of the face. This specific presentation can be described using Action Units (AUs) ^23^, which are features widely used to quantify the movements that appear on the face due to the actions of individual muscles or groups of muscles(e.g.,^14,17,19^).

Furthermore, findings from a previous study ^21^, including the association between the Face AI score and cognitive function, suggest that such characteristic facial features may also be present at rest in the early stages of cognitive decline. However, to our knowledge, no study focused on identifying specific facial characteristics at rest that may emerge during the early stages of cognitive decline.

Therefore, this study aims to investigate whether individuals in the early stages of cognitive decline, referred to in this study as individuals with MCI level, display characteristic facial features at rest. In this study, “MCI level” refers to individuals who have not been clinically diagnosed with MCI, but whose performance on cognitive screening tests falls below the established cutoff for MCI, thereby suggesting an early stage of cognitive decline. To examine specific facial features, we compared the resting-state AUs captured from facial videos of individuals with MCI level to those of HC to examine whether distinctive AU patterns are present.

If specific AU patterns associated with MCI level are identified in this study, they could serve as key features for developing a facial-information–based tool for the early detection of MCI level cognitive decline. Because facial expressions can be easily collected in daily life, such a tool could be used to screen for cognitive decline in everyday settings, and individuals who screen positive could then undergo more comprehensive evaluations at medical institutions to determine whether they meet the diagnostic criteria for MCI.

Furthermore, these findings may inform the selection and weighting of facial features in machine learning models to enhance screening accuracy. From a basic research perspective, this study may contribute to understanding the underlying mechanisms through which facial characteristics manifest in individuals with MCI level.

## Methods

### Participants

A total of 50 participants (19 males and 31 females) were recruited for the study. Inclusion criteria were: (i) age 65 years or older, and (ii) being native Japanese speaker. Exclusion criteria included: (1) use of medications that affect cognitive function, (2) a history of neurological or psychiatric disorders that may impact the central nervous system, and (3) a history of severe head trauma.

### Procedure

To comprehensively characterize MCI level across various domains, a multimodal dataset was acquired, including facial data captured during rest, conversation, and a facial expression task in which participants were instructed to voluntarily produce happy and angry facial expressions, conversational data, cognitive assessments, questionnaires, eye-tracking metrics, electroencephalography (EEG), and magnetic resonance imaging (MRI). In the present study, facial data recorded during the resting state, together with scores from the Japanese version of the Montreal Cognitive Assessment (MoCA-J) ^24^, and the Geriatric Depression Scale (GDS) ^25^, were employed to investigate facial features associated with MCI level in a non-task condition. Analyses of the remaining modalities will be reported separately to provide further insight into the multidimensional aspects of MCI level.

### Cognitive assessment

The MoCA-J, a screening tool for MCI with confirmed validity and reliability ^24^, was used. The MoCA-J is known to have higher accuracy in detecting MCI ^24^ that assesses multiple cognitive domains including memory, language, executive function, visuospatial skills, calculation, abstraction, attention, concentration, and orientation ^26^. The MoCA-J takes approximately 10 minutes to administer, and the results are scored out of 30 points. A score of 26 or above is considered HC individual, while a score of 25 or lower is considered suspicion of MCI individual ^24^. This study followed this criterion, dividing participants into MCI level groups and HC groups.

### Assessment of facial expression

Participants were told, “With your eyes open, look at the fixation cross for 1 minute, and keep your mind as clear as possible.” Their facial expressions were recorded using a web camera. To minimize noise from responses to instructions and nodding, we excluded the first and last 5 seconds of the 1-minute recording and analyzed the remaining 50 seconds of facial expressions.

Facial Action Coding System is a comprehensive, anatomically based system for describing all visually discernible facial movements ^23^. It describes facial expressions by decomposing them into AUs.

OpenFace 2.0 is an open-source tool that employs machine learning models to recognize a subset of 17 or 18 AUs, specifically AU1, 2, 4, 5, 6, 7, 9, 10, 12, 14, 15, 17, 20, 23, 25, 26, 28, and 45 from videos ^27–29^.

OpenFace 2.0 quantifies the movement of each detected AU in terms of intensity and presence. AU intensity values represent the strength of each of the 17 AUs on a continuous scale ranging from 0 to 5, while AU presence values represent whether an AU is visible on the face, with 0 denoting its absence and 1 indicating its presence. The presence values are calculated for 18 AUs, consisting of the AUs used for intensity values plus AU28 (lip suck). OpenFace’s performance has been validated in previous studies ^27,30^. Since the AU intensity and presence predictors were trained separately on different datasets, and their predictions may not always be consistent ^28^, this study used both sets of features (AU intensity and presence) and examined the consistency of the results.

The face videos recorded in this study were captured using a webcam (model: Logitech Brio 300) under artificial lighting conditions in the conference room where the experiment took place, with a resolution of 1920×1080 pixels. Each video lasted 50 seconds, with a frame rate of 25 frames per second. As a result, each video consisted of 1250 frames, with each frame containing 17 AU intensity values and 18 AU presence values.

### Geriatric Depression Scale

Because depressive symptoms may be related to facial expressions ^14^ and depressive symptoms and MCI occasionally coexist ^31^, to prevent confounding, we assessed participants for depressive symptoms by using the Japanese version of the GDS. The GDS consists of 15 questions, including “Do you feel that your life is empty?” and other questions, which are answered by choosing “yes” or “no.” Higher scores indicate greater depressive tendencies. Its validity and reliability have also been confirmed ^25^.

### Data Analysis

#### AU intensity

For each of the 17 AUs, the following method was adopted; that is, the AU intensity value was used as the response variable, and the MCI dummy variable (MCI level= 1, HC = 0) was used as the explanatory variable. Since the response variables were nested within participants, a multilevel model with random intercepts was adopted. As already noted, AU intensity ranges from 0 to 5. We divided this value by 5 to convert the range to a scale of 0 to 1 so that beta regression analysis can be applied. Beta regression analysis is not applicable when the response variable includes exactly 0 and/or exactly 1. Since the distribution of AU intensity contained unignorable amounts of zeros as shown in the Result section, a zero-inflated beta regression model was adopted. This model consists of two components. One predicted continuous values strictly between 0 and 1 (beta component), while the other predicted the likelihood of 0 occurring (zero-inflation component). Hereafter, the regression coefficients of the MCI dummy variable in each component are written as β_beta_ and β_zero_, respectively, using subscripts to identify which component the statistics refer to. Because in both components a logit link was applied to the linear predictor, the fixed effects of the MCI dummy variable are interpreted as follows: In the beta component, being MCI level is associated with a β_beta_ increase in the logit-transformed mean of AU intensity converted to a value in the range 0 to 1; in the zero-inflation component, being MCI level is associated with a β_zero_ increase in the log-odds of the AU intensity being exactly zero. For AU7 (lid tightener), the proportion of values exactly equal to 1 was 0.0036 in the MCI level group and 0 in the HC group, which was substantially lower than the proportion of values exact 0 as shown in the Result section, so for convenience, we subtracted 10^-3^ from the value when it was 1. The analysis was performed using the glmmTMB package ^32^ in R Statistical Software.

#### AU presence

For each of the 18 AUs, the following method was adopted; that is, the AU presence value was used as the response variable, and the MCI dummy variable was used as the explanatory variable. Since the response variable was binary and its observations were nested within participants, we selected a multilevel logistic regression model with random intercepts. The analyses were performed using the lme4 package ^33^ in R Statistical Software.

#### Multiple comparison correction

Bonferroni correction was applied jointly to all tests for AU intensity and AU presence. For AU intensity, the effect of the MCI dummy variable was tested in both the beta and zero-inflation components for each of the 17 AUs (34 tests in total), and for AU presence, the effect of the MCI dummy variable was tested for each of the 18 AUs (18 tests), resulting in 52 tests overall. Accordingly, the corrected significance threshold was set at 0.05/52 = 9.62 × 10^-4^.

## Result

### Demographics

Data from 5 participants were excluded from the analysis because OpenFace was unable to accurately detect eyebrow positions due to obstructions caused by eyeglasses or bangs. Thus, 31 MCI level participants and 14 HC participants were included in the analysis. Participants’ demographics are shown in Table 1. Since there was no significant difference between the MCI level and HC groups in the GDS score (*p* = 0.74) using the t-test, GDS scores were not controlled in the subsequent analysis.

**Table 1.**
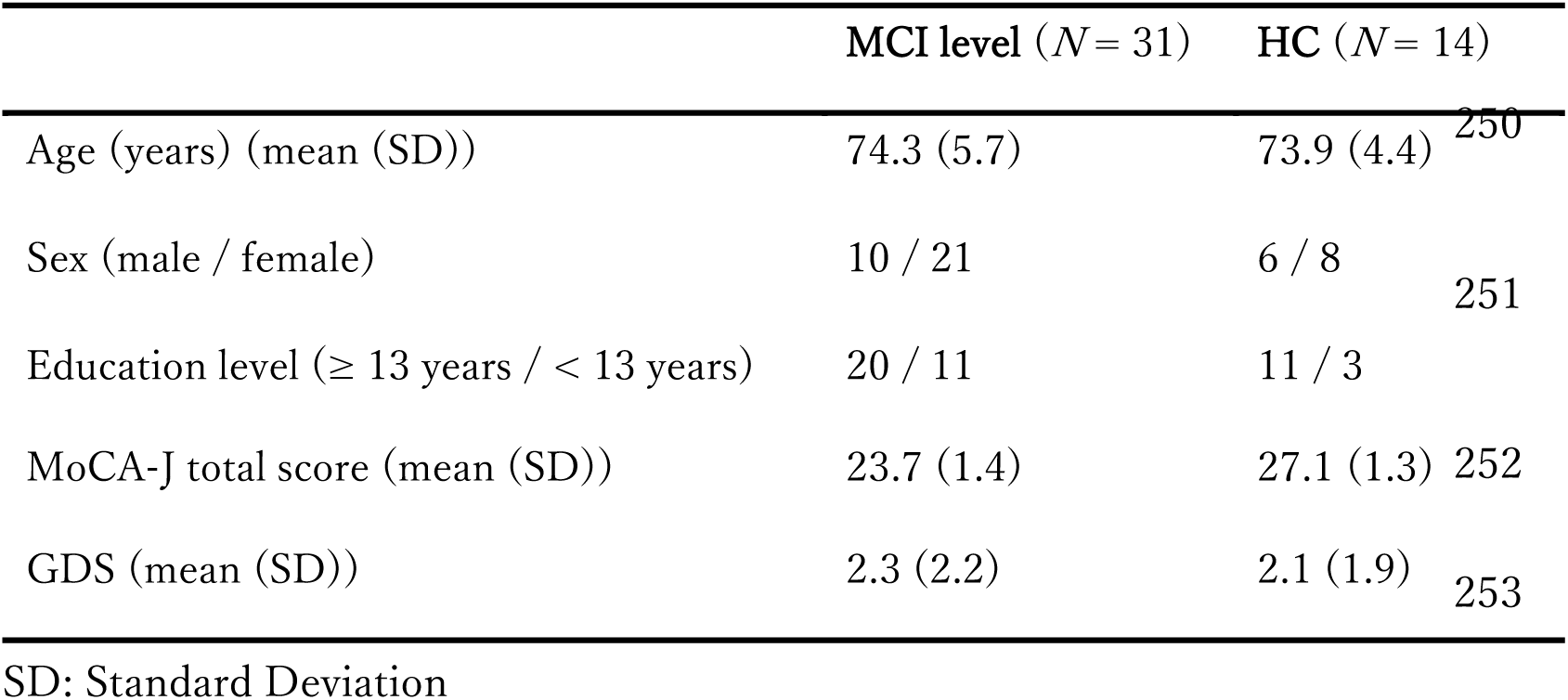
Participants’ demographics.

### AU intensity

The distribution characteristics of AU intensity are presented in Table 2. Our multilevel zero-inflated beta regression results showed that for all 17 AUs, the fixed effect of the MCI dummy variable in the beta component was not significant. For the zero-inflation component, the fixed effect of the MCI dummy variable was significant for the AU10 (upper lip raiser; β_zero_ = 9.93, SE_zero_ = 2.75, *p* < 0.001) and AU17 (chin raiser; β_zero_ = 5.39, SE_zero_ = 2.72, *p* = 0.048). The estimated variances of the random intercept were 104.60 and 56.71, respectively. Only AU10 remained significant after multiple comparisons. An additional results file shows this in greater detail [see Supplementary file].

**Table 2.** Distribution characteristics of AU intensity.

| | | MCI level ($N=31$) | | | HC ($N=14$) | | |
| --- | --- | --- | --- | --- | --- | --- | --- |
|  | FACS name | Mean | SD | Proportion<br>of zeros | Mean | SD | Proportion<br>of zeros |
| AU1 | Inner brow<br>raiser | 0.29 | 0.14 | 0.008 | 0.29 | 0.16 | 0.00006 |
| AU2 | Outer brow<br>raiser | 0.38 | 0.19 | 0.080 | 0.29 | 0.19 | 0.057 |
| AU4 | Brow<br>lowerer | 0.09 | 0.11 | 0.312 | 0.08 | 0.10 | 0.366 |
| AU5 | Upper lid raiser | 0.07 | 0.08 | 0.322 | 0.08 | 0.10 | 0.457 |
| AU6 | Cheek raiser | 0.19 | 0.11 | 0.064 | 0.21 | 0.16 | 0.046 |
| AU7 | Lid | 0.45 | 0.24 | 0.056 | 0.40 | 0.18 | 0.002 |
|  | tightener |  |  |  |  |  |  |
| AU9 | Nose | 0.10 | 0.11 | 0.290 | 0.10 | 0.10 | 0.270 |
|  | wrinkler |  |  |  |  |  |  |
| AU10 | Upper lip | 0.08 | 0.10 | 0.412 | 0.14 | 0.12 | 0.138 |
|  | raiser |  |  |  |  |  |  |
| AU12 | Lip corner | 0.10 | 0.12 | 0.355 | 0.12 | 0.11 | 0.159 |
|  | puller |  |  |  |  |  |  |
| AU14 | Dimpler | 0.11 | 0.12 | 0.354 | 0.11 | 0.14 | 0.353 |
| AU15 | Lip corner | 0.26 | 0.14 | 0.044 | 0.31 | 0.13 | 0.000 |
|  | depressor |  |  |  |  |  |  |
| AU17 | Chin raiser | 0.05 | 0.07 | 0.480 | 0.08 | 0.10 | 0.240 |
| AU20 | Lip | 0.31 | 0.16 | 0.067 | 0.29 | 0.14 | 0.024 |
|  | stretcher |  |  |  |  |  |  |
| AU23 | Lip | 0.03 | 0.04 | 0.481 | 0.04 | 0.05 | 0.428 |
|  | tightener |  |  |  |  |  |  |
| AU25 | Lips part | 0.16 | 0.10 | 0.071 | 0.12 | 0.14 | 0.208 |
| AU26 | Jaw drop | 0.17 | 0.13 | 0.145 | 0.13 | 0.14 | 0.300 |
| AU45 | Blink | 0.18 | 0.15 | 0.169 | 0.21 | 0.12 | 0.107 |

### AU presence

The distribution characteristics of AU presence are presented in Table 3. Multilevel logistic regression results showed that, of the 18 AUs, the fixed effects of the MCI dummy variable were significant for AU23 (lip tightener; β = -3.62, SE = 1.03, *p* < 0.001) and AU28 (lip suck; β = 20.36, SE = 1.42, *p* < 0.001). The estimated variances of the random intercept were 29.49 and 254.94, respectively. They were also significant after multiple comparisons. An additional results file shows this in greater detail [see Supplementary file].

**Table 3.** Distribution characteristics of AU presence.

| | | **MCI level** ($N=31$) | **HC** ($N= 14$) |
| --- | --- | --- | --- |
|  | FACS name | Proportion of zeros | Proportion of zeros |
| AU1 | Inner brow raiser | 0.093 | 0.001 |
| AU2 | Outer brow raiser | 0.139 | 0.030 |
| AU4 | Brow lowerer | 0.917 | 0.780 |
| AU5 | Upper lid raiser | 0.693 | 0.715 |
| AU6 | Cheek raiser | 0.662 | 0.523 |
| AU7 | Lid tightener | 0.418 | 0.388 |
| AU9 | Nose wrinkler | 0.999 | 0.928 |
| AU10 | Upper lip raiser | 0.829 | 0.648 |
| AU12 | Lip corner puller | 0.808 | 0.819 |
| AU14 | Dimpler | 0.637 | 0.701 |
| AU15 | Lip corner depressor | 0.113 | 0.066 |
| AU17 | Chin raiser | 0.906 | 0.705 |
| AU20 | Lip stretcher | 0.291 | 0.094 |
| AU23 | Lip tightener | 0.647 | 0.419 |
| AU25 | Lips part | 0.882 | 0.905 |
| AU26 | Jaw drop | 0.780 | 0.784 |
| AU28 | Lip suck | 0.925 | 1.000 |
| AU45 | Blink | 0.460 | 0.357 |

## Discussion

In this study, we tested the hypothesis that the facial features of MCI level at resting state appear in AUs, which refers to the appearance of how facial muscles are used. We aimed to develop a tool for the early detection of MCI level cognitive decline, using facial expressions and to obtain knowledge that will contribute to basic research on the facial expression characteristics of participants experiencing early cognitive impairment. Such an approach is important because facial-expression–based screening could provide a simple and accessible method for detecting early cognitive decline using a modality other than conventional cognitive assessments. Individuals identified through this screening could then undergo further diagnostic evaluations, such as blood tests, at medical institutions. Therefore, this approach may serve as a preliminary screening method for Alzheimer’s disease and MCI. In general, facial expression analysis for the early detection of dementia, MCI, has not been sufficiently studied ^34^, and therefore, the findings of this study, which investigated specific AU features of MCI level, are novel.

Our analysis showed that participants with MCI level were more likely than HC to have zero intensity for AU10 (upper lip raiser) (from the AU intensity data) and the absence of AU23 (lip tightener) and the presence of AU28 (lip suck) (from the AU presence data). In addition, because GDS scores did not differ significantly between MCI level group and HC groups, the observed differences in AU10, AU23 and AU28 are unlikely to be confounded by depressive symptoms. The difference observed between the results for AU intensity and presence may be because AU28 is computed based solely on the presence feature. Meanwhile, the results for AU10 and AU23 may reflect differences in the prediction models and training datasets used for estimating AU intensity and presence ^28^. It has been stated that these two types of predictions do not necessarily yield identical results ^28^.

Although the significant AUs differed across the data, they are all located near mouth. Previous studies reported facial expression differences between HC and dementia participants during neutral expressions, with particularly pronounced differences in the lower half of the face ^21^. Thus, our results extend their finding to individuals in the early stages of cognitive decline without a diagnosis of dementia.

Furthermore, the finding that AU10 and AU23 are less likely to appear in MCI level group, but AU28 is more likely to appear in MCI level group, suggests that MCI level group may not simply be characterized by consistently infrequent movement of facial muscles and a lack of facial expression, but may rather be characterized by the facial muscles used. Furthermore, our findings of infrequent movements of AU10 and AU23 and frequent movement of AU28 in MCI level group caution against relying only on infrequent movement of AUs to detect early stages of cognitive decline.

There are several possible explanations for the appearance of AU features around the mouth of MCI level participants. For example, individuals with MCI may experience difficulties in social interactions due to cognitive impairments, leading to reduced opportunities for social engagement. This reduction may result in decreased activity of the muscles around mouth, which could appear as features around the mouth during resting states. Alternatively, limited social interaction itself may be independently associated with both the onset of MCI and reduced mouth movement. Future research taking levels of social engagement into consideration may help to clarify the causality.

In addition, since facial expressions are deeply related to emotions ^35^, it is possible that differences between the HC and MCI level groups in the occurrence of emotions in daily life had an influence. For example, AU10 has been reported to be associated with smiling ^36^. Previous studies on smiling have reported differences in smile indices calculated using deep learning methods between HCs and individuals with AD or MCI ^20^. Additionally, it has been suggested that individuals with cognitive impairment, including dementia, may have a particularly low probability of expressing happiness-related emotions during conversations, compared to the HC group ^37^. These findings suggest, for example, that individuals with MCI level may have more difficulty experiencing emotions that lead to smiling in daily life compared to those in the HC group. This reduced emotional elicitation might influence the reduced activation of facial muscles associated with smiling, potentially contributing to the observed differences in facial muscle activity at rest between the MCI level and HC groups. These possibilities may be explored through the examination of indicators of emotional experience in individuals with MCI level, including self-report questionnaires and structural and functional changes in brain regions associated with emotional elicitation.

One additional possible background factor that could explain the facial expression differences in this study, which was not included in the original hypothesis, is the potential influence of stress during the experiment. For instance, AU10 and AU23, which showed differences in the present results, correspond to AUs that appear during the stress response ^38^. In this experiment, the instructions were designed to avoid eliciting emotional responses, and the participants were instructed to remain in a resting state. However, given the nature of the experimental setting, it is possible that the participants still experienced psychological stress, such as tension. One possible explanation for the reduced expression of AU10 and AU23 in the MCI level group is that the HC group experienced more stress, while the MCI level group felt less stress. Alternatively, both groups may have experienced similar levels of stress, but individuals with MCI level may have been less likely to display these specific facial AUs. In the future, the simultaneous measurement of heart rate as a stress indicator ^38^, along with assessments of participants’ subjective stress levels and group differences in stress status, may provide further insight into the factors influencing facial expressions during the resting-state task in individuals with MCI level. In contrast, if the increased expression of AU10 and AU23 in the HC group reflects a stress-related response, the MCI level group may exhibit a different pattern, characterized by a higher occurrence of AU28. One possible explanation is that, under stress, the movement of AU28 is more effectively regulated in healthy individuals, whereas such regulation may be diminished in individuals with MCI level. As a result, AU28 may be more frequently expressed in the MCI level group. This may suggest that cognitive decline is associated with altered regulation of specific facial muscle activities under stress conditions.

It may be difficult for participants to experience no stress at all in experimental settings. However, in the future, facial-based tools for the early detection of cognitive decline may be used in a wide range of situations, including low-stress environments such as everyday life. Therefore, investigating in detail the facial features that only appear under stress conditions will be an important factor in determining the appropriate contexts for using screening tools.

Despite our promising fixed effects for MCI level detection, individual differences may also play an important role and warrant consideration. Additional exploratory analyses were conducted by including age, gender, and GDS as control variables; however, the variance did not decrease. In the future, it will be important to identify additional factors that explain individual differences in facial expression features and to investigate the facial characteristics of MCI level using a more stable model that controls for the influence of these factors.

One limitation of this study is the homogeneity of the participant sample, as all individuals were Japanese. To assess the generalizability and external validity of these findings, future research should replicate the study across diverse populations and cultural contexts. In addition, a potential limitation relates to self-selection bias arising from the collection of facial biometric data. As this requirement was explicitly communicated during the participant recruitment process, individuals who were reluctant to provide such data may have opted not to participate. Consequently, these individuals are not represented in the sample, which may limit the generalizability of the findings. Additionally, attitudes toward the collection of facial biometric data may vary across cultural contexts, with some populations potentially expressing greater hesitation or concern.

### Conclusions

Characteristic facial features of individuals at the MCI level during the resting state were observed in AUs related to upper lip raiser, lip tightener, and lip suck. These findings suggest that facial movements around the mouth may indicate MCI level cognitive decline. These results may contribute to the development of facial expression-based MCI screening tools in the future.

## Consent for publication

Not applicable.

## Data availability

The datasets built and analyzed during the current study are not publicly available at the time of submission but are available from the corresponding author upon reasonable request.

## Acknowledgements

The authors would like to thank Sachiko Iwata for her crucial support in participant recruitment, which significantly contributed to the success of this research.

## Authors’ contributions

MM, KI and MOM designed this study. MM, HS, KT, AW, KK and KF collected data. KWM and MOM contributed to the ethical design of the study and managed the ethics review process. MM analyzed the data under the supervision of TS. MM wrote the manuscript under the supervision of TS, TK, SO, KI, HS and MOM. All authors have read and approved the final manuscript.

## Supplementary material

Supplementary file.pdf

Results for facial expression data.

**Table S1.** Result of multilevel zero-inflated beta regression analysis for AU intensity.

**Table S2.** Result of multilevel logistic regression analysis for AU presence.

