## Supplementary file for "Characteristic resting state facial expressions in older adults with mild cognitive impairment level"

**Table S1.** Result of multilevel zero-inflated beta regression analysis for AU intensity

| | Zero-inflated beta regression | | | | | | | |
|---|---|---|---|---|---|---|---|---|
| | Zero-inflated component | | | | beta component | | | |
| | $\beta_{zero}$ | SE | $p$ value | Variance of intercepts | $\beta_{beta}$ | SE | $p$ value | Variance of intercepts |
| AU1 | 0.78 | 4.09 | 0.85 | 128.98 | -0.02 | 0.30 | 0.95 | 0.88 |
| AU2 | -0.47 | 3.00 | 0.88 | 224.80 | 0.49 | 0.45 | 0.28 | 1.96 |
| AU4 | -2.85 | 3.85 | 0.46 | 90.52 | -0.09 | 0.41 | 0.82 | 1.29 |
| AU5 | -0.86 | 1.71 | 0.61 | 26.79 | 0.18 | 0.35 | 0.60 | 1.07 |
| AU6 | -0.19 | 3.00 | 0.95 | 238.26 | 0.02 | 0.33 | 0.95 | 1.02 |
| AU7 | 0.56 | 4.57 | 0.90 | 234.42 | 0.24 | 0.42 | 0.56 | 1.68 |
| AU9 | 0.95 | 2.45 | 0.70 | 395.85 | -0.12 | 0.41 | 0.76 | 1.38 |
| AU10 | 9.93 | 2.75 | $3.01\times10^{-4}$** | 104.60 | -0.66 | 0.42 | 0.12 | 1.62 |
| AU12 | 1.03 | 2.79 | 0.71 | 535.67 | -0.43 | 0.43 | 0.32 | 1.55 |
| AU14 | -1.82 | 3.43 | 0.60 | 82.07 | 0.08 | 0.42 | 0.85 | 1.45 |
| AU15 | 9.52 | 362.80 | 0.98 | 409.23 | -0.28 | 0.25 | 0.26 | 0.60 |
| AU17 | 5.39 | 2.72 | $4.78\times10^{-2}$* | 56.71 | -0.19 | 0.35 | 0.58 | 1.04 |
| AU20 | 0.35 | 4.65 | 0.94 | 551.10 | 0.28 | 0.23 | 0.23 | 0.51 |
| AU23 | 0.46 | 2.69 | 0.86 | 60.81 | -0.05 | 0.33 | 0.89 | 0.86 |
| AU25 | -0.67 | 2.35 | 0.78 | 250.24 | 0.60 | 0.35 | 0.08 | 1.10 |
| AU26 | -1.03 | 2.43 | 0.67 | 355.93 | 0.40 | 0.37 | 0.28 | 1.14 |
| AU45 | 0.88 | 2.72 | 0.75 | 245.59 | -0.45 | 0.29 | 0.12 | 0.77 |

One asterisk represents $p<0.05$ and two asterisks indicate significance after multiple comparisons.

SE: Standard Error

**Table S2.** Result of multileve logistic regression analysis for AU presence

| | $\beta$ | SE | $p$ value | Variance of intercepts |
|---|---|---|---|---|
| AU1 | -0.91 | 2.18 | 0.68 | 568.32 |
| AU2 | -1.64 | 2.22 | 0.46 | 845.98 |
| AU4 | -0.78 | 0.77 | 0.31 | 395.93 |
| AU5 | 0.24 | 0.91 | 0.79 | 224.12 |
| AU6 | -5.28 | 5.80 | 0.36 | 141.96 |
| AU7 | -0.43 | 1.37 | 0.75 | 1032.29 |
| AU9 | -1.40 | 3.50 | 0.69 | 357.98 |
| AU10 | -1.41 | 1.02 | 0.17 | 606.09 |
| AU12 | -0.22 | 2.35 | 0.93 | 287.25 |
| AU14 | 0.11 | 1.06 | 0.92 | 713.47 |
| AU15 | -0.23 | 1.08 | 0.83 | 114.43 |
| AU17 | -1.24 | 0.85 | 0.14 | 319.92 |
| AU20 | -0.91 | 1.18 | 0.44 | 375.73 |
| AU23 | -3.62 | 1.03 | $4.45\times10^{-4}$** | 29.49 |
| AU25 | 1.03 | 1.19 | 0.38 | 246.31 |
| AU26 | 0.57 | 2.82 | 0.84 | 330.09 |
| AU28 | 20.36 | 1.42 | $2.22\times10^{-16}$** | 254.94 |
| AU45 | -4.24 | 2.31 | 0.07 | 63.75 |

Two asterisks indicate significance after multiple comparisons.
SE: Standard Error
